# Angiographic Burden of Coronary Atherosclerosis Contributes To Adverse ASCVD Outcomes Independent Of Traditional Risk Factors

**DOI:** 10.1101/2025.04.06.25325252

**Authors:** Noah L. Tsao, Sarah A. Abramowitz, Gabrielle E. Shakt, Renae Judy, Austin T. Hilliard, Penn Medicine Biobank, Scott M. Damrauer, Themistocles L Assimes, Shoa L Clarke, Catherine Tcheandjieu, Michael G. Levin

**Affiliations:** Department of Surgery, University of Pennsylvania Perelman School of Medicine, Philadelphia, PA, USA; VA Palo Alto Health Care System, Palo Alto, CA, USA; Department of Medicine, Division of Cardiovascular Medicine, Stanford University School of Medicine, Stanford, CA, USA; Corporal Michael J. Crescenz VA Medical Center, Philadelphia, PA, USA; Cardiovascular Institute, Stanford University School of Medicine, Stanford, CA, USA; Department of Epidemiology and Population Health, Stanford University School of Medicine, Stanford, CA, USA; Gladstone Institute of Data Science and Biotechnology, Gladstone Institutes, San Francisco, CA, USA; Department of Epidemiology and Biostatistics, University of California San Francisco, San Francisco, CA, USA; Division of Cardiovascular Medicine, Department of Medicine, University of Pennsylvania Perelman School of Medicine, Philadelphia, PA, USA

**Author notes:** **Correspondence:** Michael Levin, MD, 3400 Civic Center Blvd., Smilow 11, Philadelphia, PA 19104.

## Abstract

Coronary artery disease (CAD) is a major contributor to cardiovascular morbidity (including myocardial infarction [MI] and heart failure [HF]), and mortality. Although the burden of CAD (number and degree of coronary artery stenosis) has been observationally linked to these outcomes, the causal contribution and independence from traditional cardiovascular risk factors has been poorly defined. Using publicly available genome wide association study (GWAS) data and individual level outcomes data from the Penn Medicine Biobank, we investigated whether angiographic CAD burden contributes to adverse cardiovascular outcomes independent of traditional risk factors. We developed and validated a polygenic risk score (PRS) for angiographic CAD burden, demonstrating that increasing levels of the PRS were strongly associated with increased prevalence of non-obstructive and obstructive CAD on coronary angiography, and was associated with other forms of cardiometabolic disease including peripheral artery disease and atherosclerotic risk factors including hyperlipidemia, hypercholesterolemia and hypertension. Through Mendelian randomization analyses, we found that lipid measures (ApoB, HDL, LDL, total cholesterol, triglycerides) and type 2 diabetes significantly influenced myocardial infarction risk through their effects on angiographic CAD burden. Furthermore, LDL and total cholesterol demonstrated significant indirect effects on heart failure through angiographic CAD burden, suggesting these lipids primarily influence heart failure through their impact on coronary atherosclerosis. Our findings indicate that angiographic burden of coronary atherosclerosis mediates a substantial proportion of the relationship between traditional cardiovascular risk factors and adverse outcomes. These results support prioritizing primary prevention efforts targeting modifiable risk factors to prevent development and progression of coronary plaques before clinical disease manifestation.

## BACKGROUND

Over the past 15 years genome-wide association studies have identified hundreds of risk loci for coronary artery disease, and more recently identified dozens of risk loci for angiographic measures of coronary artery disease burden.^1^ Comorbid conditions including hypertension, hyperlipidemia, diabetes mellitus and smoking represent common risk factors for developing coronary artery disease (CAD), and CAD itself remains a major cause of morbidity and mortality worldwide. Although CAD often presents with major cardiovascular events like myocardial infarction, subclinical disease represents an important risk factor for adverse cardiovascular outcomes. Epidemiologic analyses have demonstrated that the burden of coronary artery disease as measured by coronary angiography is associated with increased rates of myocardial infarction and mortality.^2^ Despite these links, guidelines have recommended withholding therapies designed to prevent the development of CAD (eg. statins) until subclinical measures of atherosclerosis are already present (Eg. elevated coronary artery calcium), based on low short-term event rates.^3^ However, how lifelong susceptibility to increased CAD burden contributes to adverse cardiovascular outcomes remains uncertain.

There are several methods to quantify the extent of coronary artery disease, including coronary artery calcium score, CT coronary angiography and invasive coronary angiography. Extensive evidence indicates that total coronary atheroma burden – the summed volume of all coronary plaques – is a major predictor of events, regardless of the presence or absence of a significant stenosis.^4,5^ Despite representing an important risk factor for adverse outcomes, quantifying CAD burden at the population level is limited by cost, complexity and potential side effects of unnecessary radiation and invasive procedures.

Genetic epidemiology approaches including polygenic risk scores (PRS) and Mendelian randomization (MR) offer the opportunity to evaluate the predictive and potentially causal roles of risk factors using genetic variants as proxies, under certain assumptions.^5^ We have previously demonstrated that genetic susceptibility to coronary artery disease in the form of a polygenic risk score associates with increased angiographic burden of disease.^1,6^ While guidelines from the American College of Cardiology and the American Heart Association emphasize cardiovascular risk assessment and pharmacologic intervention based on traditional risk factors, the extent to which angiographic CAD burden mediates the effects of these risk factors on adverse outcomes remains poorly understood. Here, we used publicly available genome wide association data of CAD burden^1^ and individual level outcomes data from the Penn Medicine Biobank (PMBB) using multivariable modeling, phenome wide association studies, and a Mendelian Randomization based mediation framework to 1) validate the association between genetic variation and angiographic burden of CAD, 2) explore the causes and consequences of increased angiographic burden of CAD, and 3) address the extent to which angiographic CAD burden mediates the effects of traditional ASCVD risk factors on cardiovascular outcomes.

## METHODS

The Penn Medicine Biobank was approved by the University of Pennsylvania Institutional Review Board and all patients enrolled in PMBB provided written, informed consent.

### Data Sources

The Penn Medicine Biobank (PMBB) is an academic healthcare system-based genomic and precision medicine cohort (> 60,000 individuals at time of data extraction) that links participant blood and tissue samples with associated health information. Procedures for recruitment, consent, data collection and genotyping have been previously described.^7^ GWAS summary statistics for angiographic CAD burden were calculated using data from the VA Million Veteran Program (MVP). The angiographic CAD burden GWAS included 41,507 individuals from MVP who underwent clinically-indicated coronary angiography, and vessels were classified as normal, non-obstructive disease, single vessel obstructive disease, double vessel obstructive disease, and triple vessel obstructive disease.^1^ GWAS summary statistics for low density lipoprotein (LDL), high density lipoprotein (HDL), total cholesterol (TC) and triglycerides (TG) were gathered from the Global Lipid Genetics Consortium (GLGC).^8^ Summary statistics for Apolipoprotein A, Apolipoprotein B and lifetime smoking index were gathered from the UK Biobank (UKBB).^9,10^ Systolic blood pressure (SBP) and diastolic blood pressure summary statistics were obtained from the International Consortium of Blood Pressure.^11^ Summary statistics for urinary albumin were obtained from Haas et al using samples from the UK Biobank.^12^ BMI summary statistics were gathered from a meta-analysis of the GIANT Consortium and UKBB.^13^ Type 2 diabetes (T2DM) summary statistics were gathered from the DIAMANTE consortium.^14^ European specific summary statistics for HBA1C were obtained from Chen et. al.^15^ The coronary artery disease (CAD) polygenic risk score was calculated using a meta-analysis from the Million Veterans Program, CARDIoGRAMplusC4D, the UK BioBank (UKBB) and Biobank Japan (BBJ)^1^ comprised of over 243,000 individuals with CAD. GWAS summary statistics for myocardial infarction were gathered from CARDIoGRAMplusC4D.^16^ Heart failure summary statistics were gathered from Levin et. al.^17^ Finally, to proxy longevity indirectly while minimizing potential effects of selection bias, we used a GWAS of parental longevity that was performed in UKBB.^18,19^ Cohort summaries can be found in the Supplemental Table 1.

### CAD burden PRS Generation

A set of PRS weights for angiographic CAD burden was generated using LDPred2, a Bayesian approach to polygenic risk scoring. The “auto” setting of LDpred2 from the ‘bigsnpr’ R package was applied to GWAS summary statistics for angiographic CAD burden from the VA Million Veteran Program, with a linkage disequilibrium (LD) reference matrix for 1,444,196 HapMap3+ variants based on European individuals.^20^ Pgsc_calc, a tool from the Polygenic Score (PGS) Catalog, was used to calculate PRSs using these weight files for individuals with imputed genotyping information in the Penn Medicine Biobank.^21^ Allele frequency differences across populations can influence PRS distributions and subsequently limit performance of raw scores.^22,23^ To mitigate the effects of allele frequency differences, downstream analysis utilized principal components analysis (PCA)-normalized scores. Scores were normalized according to pgsc_calc’s Znorm_2 method, which adjusts for differences in both mean and variance using the 1000 Genomes + HGDP reference panel.^24^ It is important to note that the calculated PRS is being used to estimate/proxy genetic liability to increased CAD burden, and no predictive modeling is reported. To validate this score, angiographic burden of CAD was extracted from coronary angiography reports in the electronic health record (EHR) using a previously described natural language processing (NLP) approach^6^ and an association analysis was performed using a Bayesian multinomial regression approach among PMBB participants to consider different qualitative levels of angiographic CAD burden (no angiographic CAD, nonobstructive (<50% stenosis or luminal irregularities, and obstructive (≥50% stenosis, stent or graft). Bayesian regression was chosen over a frequentist approach due to its advantage of being able to recover the whole range of inferential solutions, rather than a point estimate and a confidence interval as in a frequentist model.^25^

### PheWAS

A phenome-wide association study (PheWAS) was performed to identify clinical diagnoses associated with the angiographic CAD burden PRS. Briefly, international classification disease codes obtained from the electronic health record were mapped to “Phecodes,” and individuals were assigned case/control status or excluded using default mapping parameters.^26^ Associations between the angiographic CAD burden PRS and phenotypes was tested using logistic regression, adjusted for age, sex and four genetic principal components. PheWAS analysis was performed first in a stratified manner by genetically identified ancestry (African and European), as well as a transethnic analysis was performed.

### Mendelian Randomization

Mendelian randomization is a method that uses genetic variants as instrumental variables to estimate associations between modifiable risk factors and outcomes. In comparison with traditional observational study designs, MR may be less biased by residual confounding or reverse causality, as the genetic variants associated with exposures are randomly allocated at conception (independently from other risk factors), and prior to the development of the outcomes of interest. In order to ensure proper reporting for studies using Mendelian randomization, the STROBE-MR checklist was used to guide the studies described. Mendelian randomization studies use genetic variation as an instrumental variable (IV) and must fulfill instrumental variable assumptions. Applied to Mendelian randomization, these assumptions are that (i) the genotype is associated with the exposure; (ii) the genotype is associated with the outcome through the studied exposure only (exclusion restriction assumption); and (iii) the genotype is independent of other factors which affect the outcome (independence assumption).^5,27^

We constructed genetic instruments for ASCVD risk factors and angiographic CAD burden using summary data from large GWAS **(Supplemental Table 1)**. Individual study descriptive statistics and participating biobanks for each GWAS are presented in the supplemental table.

The total effect of traditional ASCVD risk factors (LDL, HDL, Total Cholesterol, Triglycerides, ApoA1, ApoB, SBP, DBP, Urinary Albumin, BMI, Type 2 Diabetes, HbA1C, Lifetime Smoking Index) on angiographic CAD burden was estimated using random-effects inverse-variance–weighted MR within the TwoSampleMR package in R. Instruments for each of the phenotypes above were created using genetic variants and their corresponding weights that were determined by previous GWAS analysis. To satisfy the first Mendelian randomization assumption, genetic variants were selected as instruments based on a genome-wide significant p-value threshold of 5×10^−8^, and clumped into approximately independent signals based on the default r^2^ threshold of 0.001 and distance threshold of 10,000kb. We further calculated F-statistics for each instrument to assess the strength of association between each instrument and the corresponding exposure. Genetic instruments with F-statistics >10 are thought to be less susceptible to weak-instrument bias.^5^ Our primary MR analyses utilized an inverse variance weighted estimator using multiplicative random effects. We performed sensitivity analyses using a weighted median estimator, as this method makes different assumptions about the presence of pleiotropic genetic variants in comparison to the primary inverse-variance weighted approach. Horizontal pleiotropy and heterogeneity (via Cochrane’s Q) were assessed using MR-Egger.^28^ MR-Egger is a method that can assess whether genetic variants have pleiotropic effects on the outcome that differ on average from zero under the instrument strength independent of direct effect (InSIDE) assumption. In this method and under the InSIDE assumption, the intercept from the MR-Egger analysis can be interpreted as the average pleiotropic effect of a genetic variant included in the analysis. ^28^ Therefore, if the intercept of the MR-Egger analysis is not equal to zero, then either the average pleiotropic effect differs from zero (known as directional pleiotropy) or the InSIDE assumption in violated (or both), providing an assessment of the validity of the instrumental variable assumptions. ^28^ Based on this approach, a non-zero intercept indicates that the inverse variance weighted estimate is biased.

To fulfill the assumptions of mediation using Mendelian randomization, reverse-MR was performed to rule-out reverse-causal associations. As a positive control, the effect of ASCVD risk factors mentioned in the previous step on CAD was estimated using random-effects inverse-variance–weighted MR. The effect of angiographic CAD burden on ASCVD Events (HF, Longevity, MI) was estimated using random-effects inverse-variance–weighted MR.

Multivariable MR was used to estimate the direct effects of the traditional ASCVD risk factors on ASCVD outcomes, and the indirect (mediating) effects of angiographic CAD burden on the ASCVD outcomes (HF, Longevity and MI).

To evaluate the contribution of each variable to the outcome in question, MR-BMA, a multivariable MR method that uses Bayesian model averaging to prioritize putative causal risk factors in situations where candidate risk factors are highly correlated, was used.

### Statistical Analysis

All analyses we performed in R version 4.2.3. (R Foundation for Statistical Computing, Vienna, Austria). PheWAS analyses were performed using the default setting in the *PheWAS* package in R. Mendelian Randomization analyses were performed using the TwoSampleMR package in R and MR-BMA analyses were performed using the *mrbma* package in R. To evaluate the association between the CAD burden PRS and angiographic burden of CAD in PMBB, Bayesian multinomial regression was performed, using “normal” coronary arteries as the reference level. For this Bayesian analysis the 95% credible interval was used to evaluate statistical significance, with a probability >95% considered important. Experiment wide significance for Mendelian randomization analyses was defined by an FDR corrected p-value value < 0.05 and Bonferroni corrected P < 0.05 for PheWAS analysis. The study was not pre-registered.

## RESULTS

### Cohort Demographics

Of the 63,104 individuals enrolled in the Penn Medicine Biobank, at the time of data extraction, there were 41,660 individuals with genotype and phenotype data available. Among the analytic cohort, the median [IQR] age was 63.2 [50.7-75.8] years, 21,114 (50.7%) were male and 29,170 (70.0 %) were genetically similar to a European reference population with the other 12,490 (30%) being genetically similar to an African reference population.

### A Polygenic Risk Score Associates with Angiographic Burden of CAD

We first sought to validate that previously identified genetic variants associated with angiographic burden of CAD in MVP were also associated with this phenotype in PMBB. Polygenic risk score weights for angiographic CAD burden were generated using summary GWAS data from Tcheandjieu et al. ^1^ and risk scores were calculated for individuals in PMBB. Ultimately, 1,406,831 genetic variants were included in the PRS. To validate the polygenic risk score, we investigated whether our genome-wide PRS was associated with extent of angiographic coronary disease (no angiographic CAD, nonobstructive (<50% stenosis or luminal irregularities, and obstructive (≥50% stenosis, stent or graft) among PMBB participants who had undergone coronary angiography (n = 3771). There were 840 individuals with angiograms rated to be normal, 840 individuals with angiograms demonstrating non-obstructive disease and 2091 individuals demonstrating obstructive disease. The median PRS was significantly greater among the obstructive group compared with the normal (p = 1.63×10^−16^) and non-obstructive (2.15×10^−12^) groups, with overlap in scores between all three groups **(Supplemental Figure 1)**. The proportion of individuals with obstructive disease in the major coronary vessels increased with increasing deciles of the PRS **(Figure 1)**.

**Figure 1.**
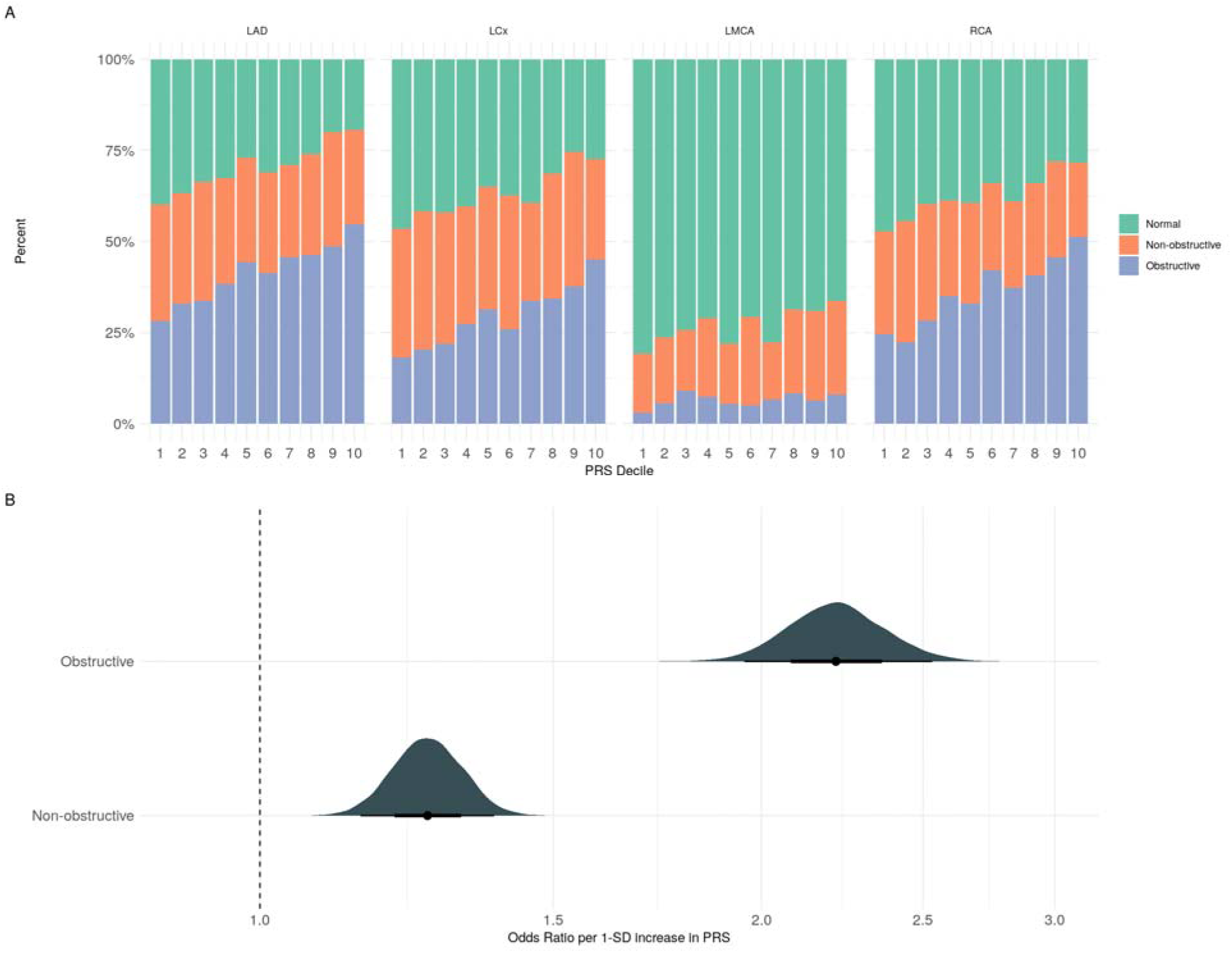
A) Distribution of angiographic CAD burden by PRS decile and vessel. The *x*-axis represents the PRS decile and *y-*axis represents percent of patients in each angiographic category that make up the PRS decile. Green represents angiographically normal individuals, orange represents individuals with angiographic disease that is non-obstructive, and purple represents individuals with angiographic disease that is obstructive. **B) Bayesian distribution of odds ratio of non-obstructive and obstructive disease per 1-SD increase in the Angiographic CAD burden PRS**. (LAD = Left Anterior Descending Artery, LCx = Left Circumflex Artery, LCMA = Left Main Coronary Artery, RCA = Right Coronary Artery)

To quantify the association between the PRS and angiographic burden of CAD, Bayesian multinomial logistic regression was performed. Increasing levels of the PRS were associated with increased prevalence of non-obstructive (odds ratio of 1.26 per 1 SD increase in the PRS for non-obstructive disease as compared to individuals with no angiographic disease [95% credible interval 1.14-1.39]) and obstructive CAD (OR 2.23 per 1 SD increase in the PRS for obstructive disease as compared to individuals with no angiographic disease [95% credible interval 1.94-2.55]). This result suggests that our polygenic risk score is measuring genetic risk of increased angiographic burden of CAD in our population of interest.

### PheWAS Identifies Diseases Associated with Angiographic Burden of CAD

We next aimed to evaluate the consequences of increased angiographic CAD burden using the CAD burden PRS. To do so, we performed a phenome-wide association study in the Penn Medicine Biobank both overall and stratified by population subgroup (European and African) **(Supplemental Table 2-4)**. The angiographic CAD burden PRS was most strongly associated with phenotypes related to coronary artery disease, including coronary atherosclerosis, ischemic heart disease, and myocardial infarction, furthering our confidence that we are capturing coronary artery disease burden using our risk score. Beyond CAD, our angiographic CAD burden risk score demonstrated phenome-wide significant associations with other forms of atherosclerosis including peripheral artery disease (OR 1.17, [95% CI 1.13-1.24], P = 1.61×10^−11^), atherosclerotic risk factors including hyperlipidemia (OR 1.12, [95% CI 1.10-1.15], P = 1.46×10^−24^), hypercholesterolemia (OR 1.07, [95% CI 1.04-1.10], P = 2.6×10^−8^), and hypertension (OR 1.06, [95% CI 1.04-1.09], P = 5.3×10^−4^). The associations between the PRS and obesity (OR 1.0, [95% CI 0.98-1.02], P = 0.991) or tobacco use disorder (OR 1.06, [95% CI 0.98-1.03], P = 0.685) did not reach statistical significance. We replicated a previously demonstrated epidemiologic association of the angiographic CAD burden PRS with type 2 diabetes (OR 1.06, [95% CI 1.03-1.08], P = 1.17×10^−5^) (**Figure 2)**.^29,30^ These PheWAS results validate our risk score is measuring genetic risk to increased angiographic coronary artery disease burden and replicates previously demonstrated associations with atherosclerosis and common atherosclerotic risk factors.

**Figure 2.**
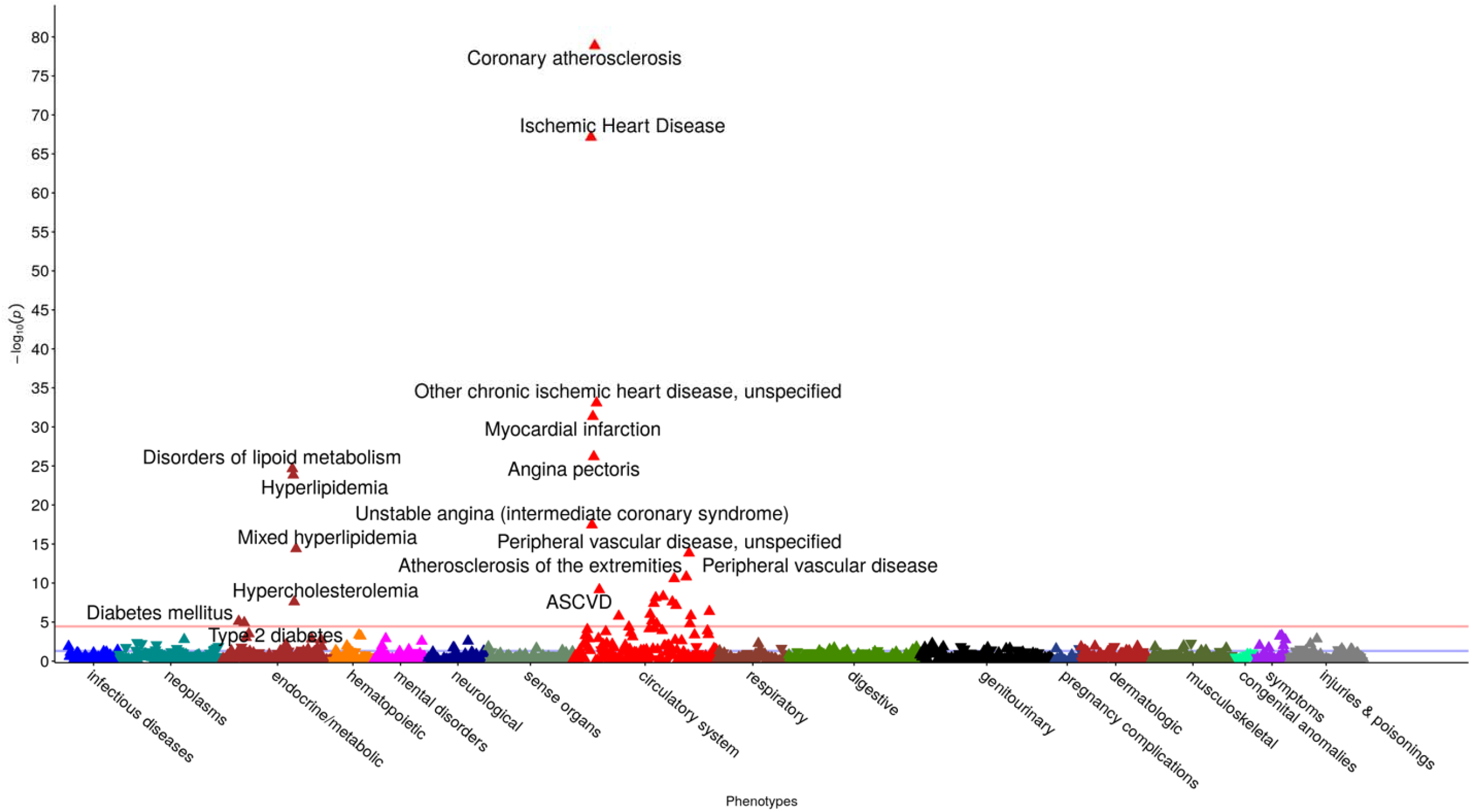
PheWAS Plot of phenotype associations with the angiographic CAD burden PRS. Results of multi-ancestry PheWAS of the angiographic CAD-burden PRS using a logistic regression model. A PheWAS Manhattan plot of phenome-wide significant (p < 3.6E-05) associations. Each point represents a phenotype. The *x-*axis represents all phenotypes, and the *y-*axis represents the strength of association as represented by −log_10_(*p* value)

### Associations Between Traditional Risk Factors and Increased Angiographic CAD burden

We next sought to quantify the associations between traditional cardiovascular risk factors and increased burden of CAD using Mendelian randomization. Prior to conducting Mendelian randomization analyses we assessed the validity of our instruments via the F statistic. The median F statistics for each instrument ranged from 36.3 to 52.6 with none demonstrating an F statistic less than 10 indicating the level of weak instrument bias is likely to be small (**Supplemental Table 5)**. We focused broadly on a set of traditional risk factors for atherosclerosis and used a Mendelian randomization framework to first confirm their associations with CAD as defined by Tcheandjieu et. al.^1^ Each risk factor except for urinary albumin demonstrated experiment-wide significant causal associations for 13 separate exposure phenotypes **(Supplemental Table 6)**.

We then sought to understand if genetic liability to these traditional risk factors similarly associated with increased angiographic burden of CAD. Total cholesterol, LDL-C, HDL-C, ApoB, triglycerides, type 2 diabetes, BMI, DBP, SBP, and ApoA1 demonstrated statistically significant associations with increased angiographic CAD burden [(OR 1.45, [95% CI 1.45-1.55]), (OR 1.49, [95% CI 1.44.-1.54]), (OR 0.83, [95% CI 0.80-0.85]), (OR 1.36, [95% CI 1.31-1.41]), (OR 1.34, [95% CI 1.29-1.39]), (OR 1.15, [95% CI 1.12-1.18]), (OR 1.12, [95% CI 1.07-1.18]), (OR 1.02, [95% CI 1.02-1.03]), (OR 1.01, [95% CI 1.01-1.02]), (OR 0.89, [95% CI 0.86-0.93]), respectively] **(Figure 3 and Supplemental Table 7**). These associations were consistent when assessed with a weighted median estimator **(Supplemental Table 7)**. Importantly, total cholesterol (intercept = −1.98×10^−3^, p = 2.17×10^−4^) and triglycerides (intercept = 0.00176, p = 0.00149) demonstrated evidence of experiment-wide significant directional pleiotropy based on the MR-Egger intercept test and all instruments except for Urinary Albumin, HbA1C and Lifetime Smoking index demonstrated statistically significant Cochran Q values for heterogeneity **(Supplemental Table 7)**. These results indicate the traditional risk factors influence angiographic burden of CAD with the expected direction of effect, but with potentially pleiotropic effects via total cholesterol and triglycerides.

**Figure 3.**
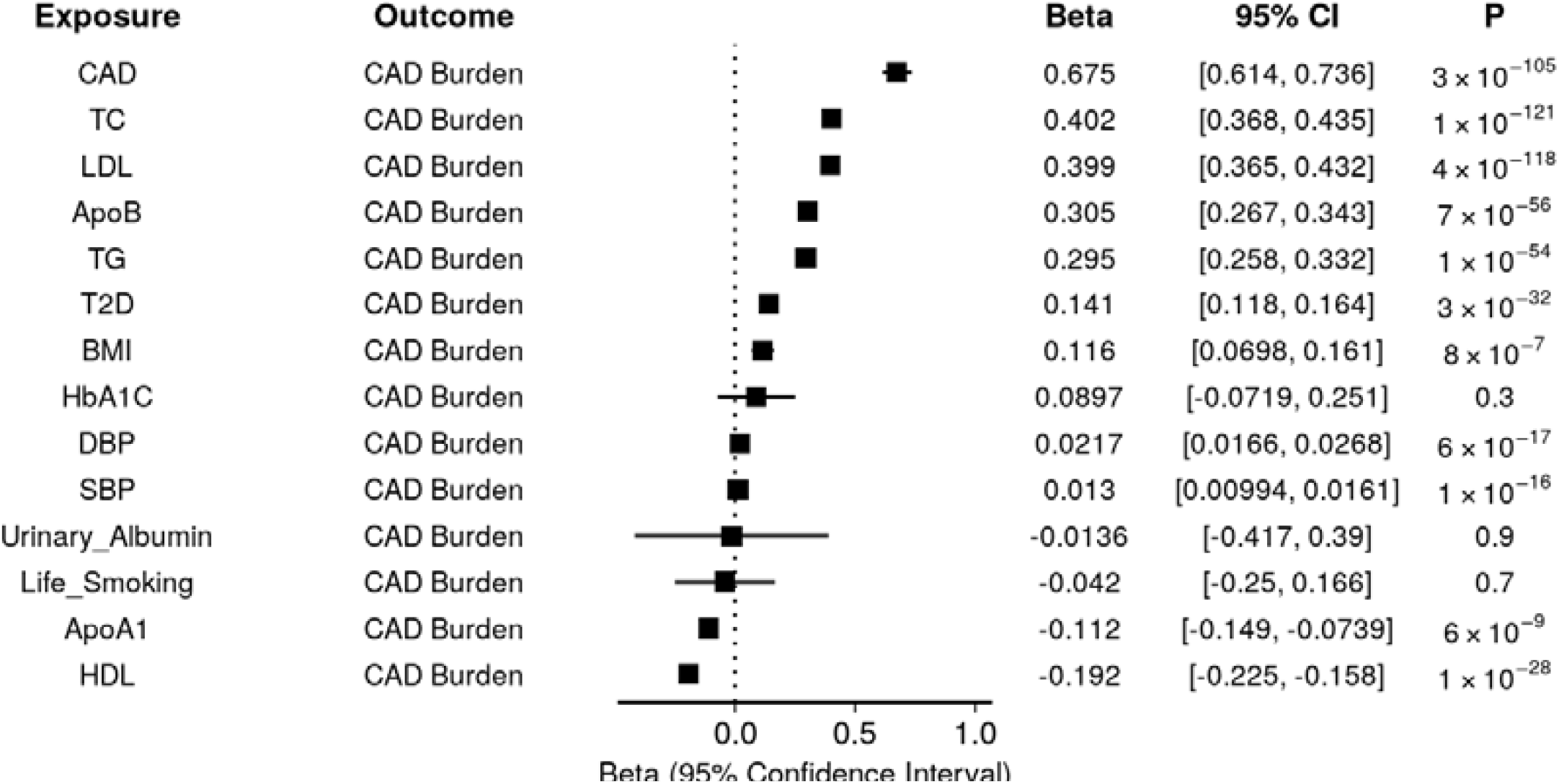
Forest Plot of MR Causal Associations of Traditional CAD Risk Factors with CAD burden. Two-sample Mendelian randomization was performed using a genetic instrument associated with CAD and traditional ASCVD risk factors on genetic liability to angiographic CAD Burden. Inverse-variance weighting was used as the primary analysis and was further examined with multiple sensitivity analyses. The Beta, 95% CIs, and *P* values are displayed.

To assess for potential bidirectional associations, we performed a reverse-MR study, in which we evaluated whether angiographic CAD burden influenced levels of traditional CAD risk factors. Angiographic CAD burden did not demonstrate experiment wide significant associations with any risk factors **(Supplemental Table 8)**. Therefore, in conjunction with the prior results, these findings suggest a unidirectional relationship in which genetic liability to traditional CAD risk factors is associated with genetic liability to angiographic CAD burden.

Because traditional risk factors for CAD often co-occur and are highly correlated, we next performed multivariable MR modeling using MR-BMA,^31^ a statistical approach to prioritize the most proximate risk factors among a set of highly correlated exposures. Although sample overlap between exposure and outcome is minimal, as noted in Supplemental Table 1, it is important to note that minimal bias is introduced by sample overlap as calculated by Burges et. al. (**Supplemental Table 9)**.^32^ In the MR-BMA analysis, type 2 diabetes was the top ranked risk factor for angiographic CAD burden (marginal inclusion probability, 0.99, *P* = 0.01). Other prioritized risk factors included SBP (marginal inclusion probability, 0.68, *P* = 0.01), HDL (marginal inclusion probability, 0.65, *P* = 0.01), total cholesterol (marginal inclusion probability, 0.61, *P* = 0.02), ApoB (marginal inclusion probability, 0.53, *P* = 0.02), and LDL (marginal inclusion probability 0.26, *P* = 0.02) **(Supplemental Tables 10-11)**. These results provide support for roles of T2D, SBP, plasma lipids and lipoproteins as primary risk factors for angiographic CAD burden.

### The Relationship between Traditional ASCVD Risk Factors and Adverse Cardiovascular Outcomes is in part Mediated by Angiographic CAD Burden

CAD has several well-established complications, including myocardial infarction, heart failure, and decreased lifespan.^33^ We sought to determine whether these adverse outcomes occur directly due to traditional risk factors themselves, or whether they are mediated by the development of coronary atherosclerosis.

To answer this question, we performed mediation analysis using multivariable Mendelian randomization, which allows us to separate direct effects (risk factor → outcome) from indirect effects (risk factor → CAD burden → outcome). Before conducting mediation analysis, we confirmed two key assumptions: (1) that our risk factors (lipids, blood pressure, etc.) were associated with the outcomes, and (2) that our proposed mediator (angiographic CAD burden) was associated with the outcomes. Both assumptions were verified **(Supplemental Table 12)**.

For myocardial infarction, we found both direct and indirect pathways. For example, BMI, blood pressure (SBP, DBP), and lipids (LDL, HDL, TC, TG) demonstrated significant direct effects on MI. However, lipid measures (ApoB, HDL, LDL, TC, TG) and type 2 diabetes also showed significant indirect effects through angiographic CAD burden **(Figure 4A, Supplemental Table 13)**. This result indicates that lipid abnormalities contribute to MI both directly and by promoting coronary atherosclerosis.

**Figure 4.**
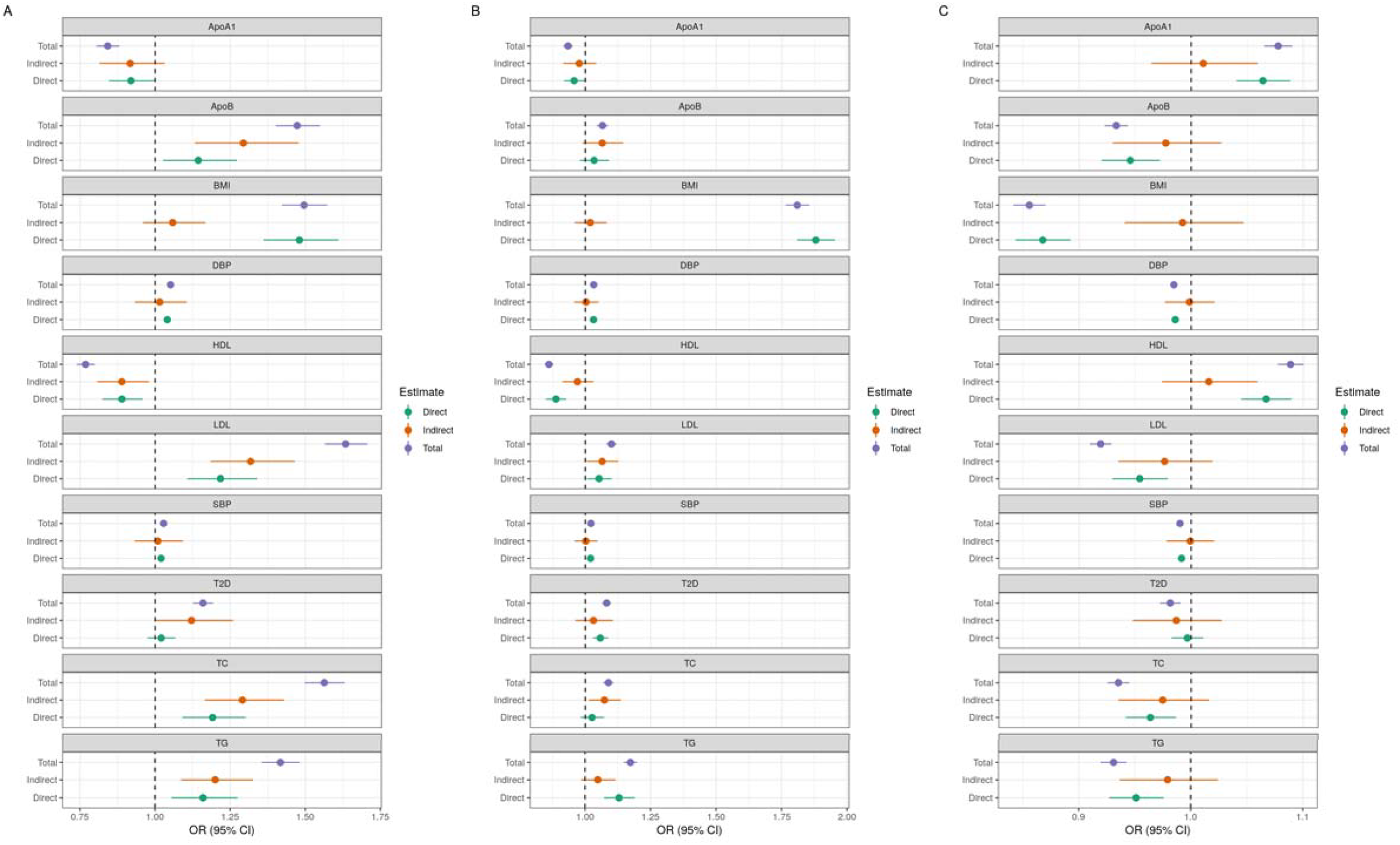
A) Association of traditional ASCVD risk factors with A) HF, B) MI, C) Longevity as mediated through angiographic CAD burden via MVMR. OR per 1-SD change in the exposure, and 95% CIs are displayed. *X-*axis represents OR (95% CI), *y-*axis demonstrates total (purple), indirect (orange) and direct (green) effects. Plots are organized by phenotype with phenotype noted in the title.

For heart failure, a different pattern emerged. Obesity (BMI), hypertension (SBP, DBP), certain lipids (HDL, TG), and type 2 diabetes had significant direct effects on heart failure. In contrast, LDL and total cholesterol influenced heart failure primarily through their indirect effects on angiographic CAD burden **(Figure 4B, Supplemental Table 13)**. This result suggests that LDL and total cholesterol contribute to heart failure mainly by promoting coronary atherosclerosis rather than through direct cardiac effects. Further analysis confirmed this pathway, showing that CAD burden primarily affects heart failure through myocardial infarction rather than directly **(Supplemental Table 14)**.

Lastly, we explored whether traditional CAD risk factors had an impact on longevity directly, or whether the effects were mediated via angiographic CAD burden. Nearly all traditional risk factors (SBP, DBP, BMI, HDL, ApoA1, ApoB, TG, LDL, TC) demonstrated significant direct effects, with the exception of type 2 diabetes. Interestingly, no risk factors showed significant indirect effects on longevity through angiographic CAD burden (Figure 4C, **Supplemental Table 13)**, suggesting that these factors influence lifespan through multiple pathways beyond just coronary atherosclerosis.

## DISCUSSION

Observational studies have linked increased angiographic CAD to worse cardiovascular outcomes, but the causal relationships between traditional risk factors, coronary atherosclerosis, and adverse outcomes have remained poorly defined. Using genetic approaches, we demonstrated that lifelong genetic susceptibility to increased angiographic CAD burden was strongly associated with adverse cardiovascular outcomes. Through mediation analysis, we found that angiographic CAD burden mediates a substantial proportion of the relationship between traditional risk factors and cardiovascular complications, particularly for lipid measures. These findings highlight several important implications: First, distinct mechanisms exist through which traditional risk factors contribute to different cardiovascular outcomes, with some pathways critically dependent on coronary atherosclerosis; Second, current guidelines that recommend waiting for CAD to develop before initiating aggressive preventive therapies may miss critical primordial and primary prevention windows for intervention; Finally, deeply phenotyped cohorts provide valuable insights into disease mechanisms that may not be captured in larger but less detailed datasets.

Our genetic evidence reveals distinct mechanisms through which traditional risk factors contribute to myocardial infarction and heart failure. For MI, we found both direct and indirect causal pathways, with lipid measures (ApoB, LDL, TC, TG) showing significant effects through both routes. This dual mechanism suggests that lipids promote MI both by increasing coronary atherosclerosis and through other pathways such as thrombogenicity and endothelial dysfunction.^34^ In contrast, for heart failure, the pathways are more differentiated by risk factor type. The significant indirect effects of LDL and total cholesterol on heart failure through angiographic CAD burden, coupled with the absence of significant direct effects, align with trials showing that statin therapy does not benefit patients with non-ischemic cardiomyopathy.^35^ This suggests that LDL’s role in heart failure is primarily related to promoting coronary atherosclerosis rather than direct myocardial effects. In contrast, the significant direct effects of obesity, hypertension, type 2 diabetes, and markers of insulin resistance (HDL, TG) on heart failure support well-established pathways for diabetic and hypertensive cardiomyopathy that operate independently of coronary atherosclerosis.^36–39^

The strong mediating role of angiographic CAD burden in the relationship between risk factors and adverse outcomes emphasizes the critical importance of prevention before coronary atherosclerosis develops. While our genetic approach cannot determine the optimal timing of interventions, these findings suggest that disrupting the development of CAD is key to preventing the adverse consequences of traditional CAD risk factors. Some current guidelines advocate for the use of coronary artery calcium scoring to guide initiation of preventive therapies, effectively waiting until CAD has already developed.^3^ Our findings suggest this approach may be suboptimal, as it allows atherosclerosis to develop before intervention. While low-short term cardiovascular event rates are used to justify withholding preventive therapies among individuals without subclinical disease, our genetic findings, which consider the lifelong genetic liability to increased CAD burden, suggest that more aggressive upstream primordial and primary prevention strategies targeting risk factors before plaque development may provide important benefit.

Our findings demonstrate that PRS can proxy angiographic CAD, consistent with prior studies.^6,40,41^ We note that the angiographic CAD burden PRS is more strongly associated with angiographic CAD than a PRS constructed from the CARDIoGRAMPlusC4D GWAS of CAD tested in Penn Medicine Biobank (OR of 2.23 as compared to 1.8 for obstructive CAD).^6^ This is consistent with the idea that deeply phenotyping a small number of individuals (e.g. 41,507 individuals from MVP who were phenotyped for angiographic CAD burden) may provide more specific association signals than larger GWAS of case-control phenotypes (e.g. 60,801 CAD cases and 123,504 controls) from CARDIoGRAMPlusC4D).^42^ Although we focused here on studying angiographic CAD burden as an endophenotype of CAD, the results here fit within the context of a larger cardiovascular genetic epidemiology literature demonstrating that genetic associations identified in smaller, well-phenotyped cohorts can provide important information about the genetic architecture of common cardiovascular traits and diseases.^43,44^

Our findings demonstrate the significant value of deeply phenotyped cohorts in understanding complex disease mechanisms. We observed that the angiographic CAD burden PRS derived from a smaller but intensively phenotyped cohort (41,507 individuals from MVP with detailed coronary angiography data) showed stronger associations with obstructive CAD (OR 2.23) compared to previous scores derived from larger case-control studies like CARDIoGRAMPlusC4D (OR 1.8).^6^ This observation highlights an important principle in genetic epidemiology: detailed phenotyping in moderate-sized cohorts can provide more specific genetic insights than larger studies with less granular phenotyping. By capturing the actual burden of coronary atherosclerosis rather than just the presence or absence of clinical CAD, we were able to more precisely dissect the pathways through which risk factors contribute to adverse outcomes. Although we focused here on studying angiographic CAD burden as an endophenotype of CAD, the results here align with recent successful genetic discovery effors in smaller, well-phenotyped cohorts that have provided important information about the genetic architecture of common cardiovascular traits and diseases.^43,44^ As we move toward increasingly personalized approaches to cardiovascular disease prevention and treatment, the value of deeply phenotyped cohorts will likely continue to grow.

This study has several limitations. First, the data collected for the PRS and PheWAS analyses come from an EHR-linked genomic and precision medicine cohort and does not include adjudicated disease status or outcomes which may have resulted in some degree of phenotype misclassification. The polygenic risk score in this study was limited to HapMap3 variants and did not examine the role of rare genetic variants known to affect angiographic CAD burden.^45^ A further limitation is that there is sample overlap between the GWAS of traditional risk factors,and the GWAS of angiographic CAD burden, although calculated bias due to this sample overlap is minimal.^32^ It is also important to note that some of the cohorts used to develop the PRS and genetic instruments are mainly hospital-based cohorts and future study is warranted to understand the generalizability of these findings to the general population.

Overall, we demonstrated that a polygenic risk score for coronary artery disease burden was associated with cardiovascular risk factors and outcomes. We observed that increased atherosclerotic plaque burden mediates a large proportion of the relationship between CAD risk factors and adverse outcomes, underscoring the role for primordial and primary prevention to prevent the development of atherosclerotic plaques.

## Data Availability

Raw data for the analysis dataset are not publicly available to preserve individuals privacy per the Health Insurance Portability and Accountability Act Privacy Rule.

