## Supplemental Figure for "Angiographic Burden of Coronary Atherosclerosis Contributes To Adverse ASCVD Outcomes Independent Of Traditional Risk Factors"

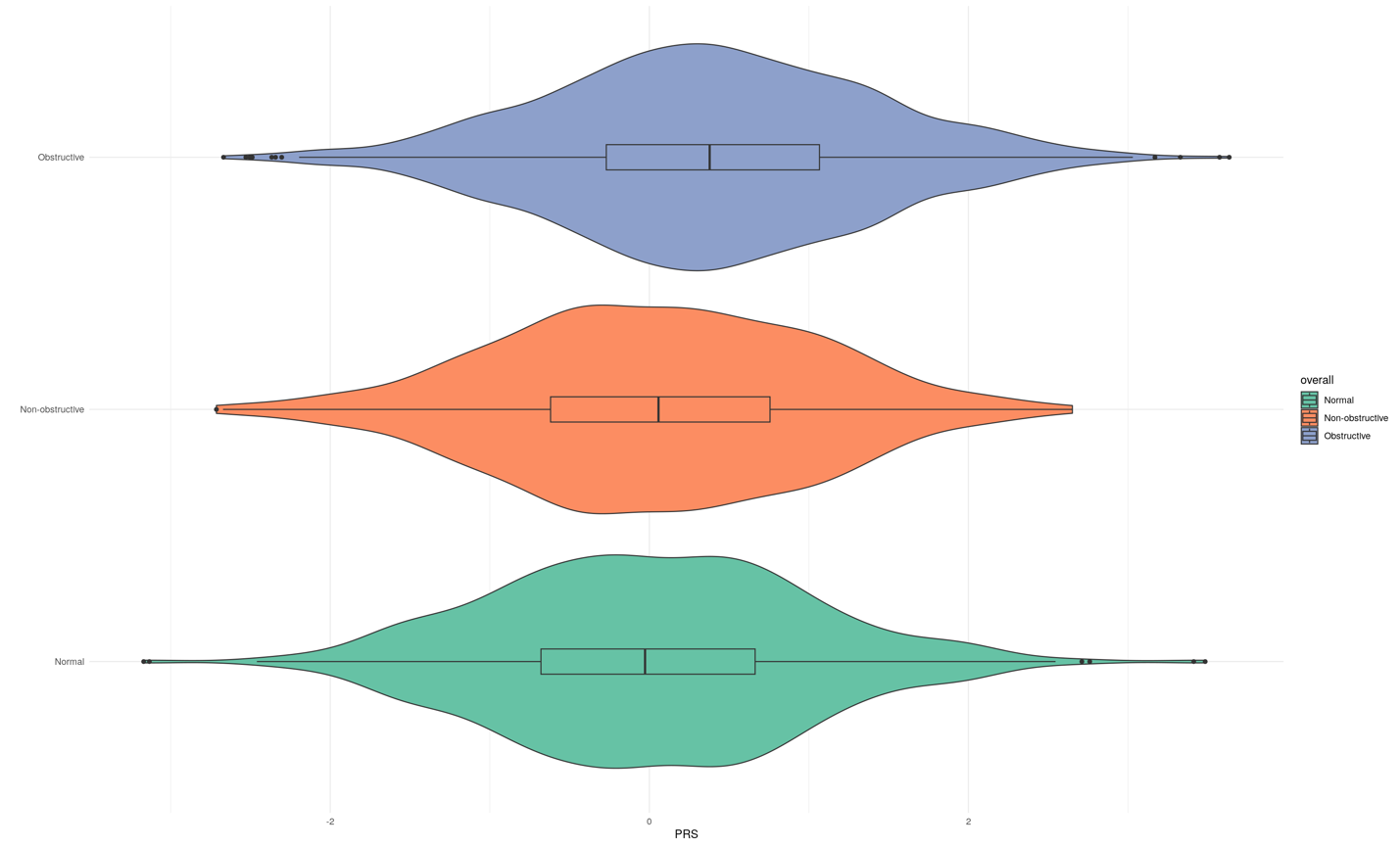

2

PRS

0

-2

Obstructive

Normal

Non-Obstructive

**Supplemental Figure 1. Violin plots demonstrating median distribution of PRS Across CAD Angiographic burden groups.** Vertical line demonstrates the median, box demonstrates the IQR.
